# Estimating the cost-of-illness associated with the COVID-19 outbreak in China from January to March 2020

**DOI:** 10.1101/2020.05.15.20102863

**Authors:** Huajie Jin, Haiyin Wang, Xiao Li, Weiwei Zheng, Shanke Ye, Sheng Zhang, Jiahui Zhou, Mark Pennington

**Author notes:** Correspondence to: Dr Huajie Jin, King’s Health Economics, Institute of Psychiatry, Psychology & Neuroscience at King’s College London, Box 024, The David Goldberg Centre, London, UK, SE5 8AF. Contributed equally (Huajie Jin, Haiyin Wang).

## Abstract

**Background:** COVID-19, an infectious disease caused by severe acute respiratory syndrome coronavirus 2 (SARS-CoV-2), swept through China in 2019–2020, with over 80,000 confirmed cases reported by end of March 2020. This study estimates the economic burden of COVID-19 in 31 provincial-level administrative regions in China between January and March 2020.

**Methods:** The healthcare and societal cost of COVID-19 was estimated using bottom-up approach. The main cost components included identification, diagnosis and treatment of COVID-19, compulsory quarantine and productivity losses for all affected residents in China during the study period. Input data were obtained from government reports, clinical guidelines, and other published literature. The primary outcomes were total health and societal costs. Costs were reported in both RMB and USD (2019 value).

**Outcomes:** The total estimated healthcare and societal cost associated with the outbreak is 4.26 billion RMB (0.62 billion USD) and 2,647 billion RMB (383 billion USD), respectively. The main components of routine healthcare costs are inpatient care (41.0%) and medicines (30.9%). The main component of societal costs is productivity losses (99.8%). Hubei province incurred the highest healthcare cost (83.2%) whilst Guangdong province incurred the highest societal cost (14.6%).

**Interpretation:** This review highlights a large economic burden of the recent COVID-19 outbreak in China. These findings will aid policy makers in making informed decisions about prevention and control measures for COVID-19.

**Funding:** The author(s) received no financial support for the research, authorship, and/or publication of this article.

**Research in context:** *Evidence before this study:* We searched MEDLINE, EMBASE and Global Health on April 4^th^, 2020, using key words and medical subject headings including (“coronavirus” OR “SARS-CoV-2” OR “COVID-19”) AND (“cost” OR “Economics” OR “resource” OR “productivity loss”). No restrictions on language or publication dates were applied. Our search did not identify any studies which reported the cost of COVID-19. Cost of severe acute respiratory syndrome (SARS), which is an infectious disease caused by another type of coronavirus – the SARS coronavirus – has been assessed by seven studies. The reported healthcare cost of managing SARS per case ranged from $4,151 USD in mainland China to $362,700 USD in Canada. The total healthcare cost and societal cost of the 2013 SARS outbreak in China took up 0.20% and 1.05% of China’s GDP, respectively. The global cost of SARS was estimated to be US $40 billion, the majority of which was caused by reduced consumer demand for goods and services due to fear associated with SARS. Two studies reported a reduction in total healthcare resource use during the peak of the SARS epidemic in Taiwan, due to people’s fears of SARS.

*Added value of this study:* To our knowledge, this study presents the first cost-of-illness study of COVID-19. The main cost components considered include identification, diagnosis, treatment and follow-up of COVID-19, compulsory quarantine and productivity loss for all affected residents in China during the study period. The total societal cost of COVID-19 was estimated to be 383 billion US dollars, which is equivalent to 2.7% of China’s gross domestic product (GDP) in 2019. Healthcare costs accounted for only 0.2% of societal cost for COVID-19, while productivity losses accounted for 99.8%. The majority of productivity losses (99.7%) were attributable to people who were not considered to have had COVID-19 but experienced lost working time due to the government policies in controlling population movement.

*Implications of all the available evidence:* Our findings suggest that the cost of COVID-19 is much larger than the cost of SARS or MERS. Productivity losses far exceeded the healthcare cost of managing COVID-19 patients. Future research is urgently required on the cost-effectiveness of different control measures of COVID-19 (e.g. policies regarding reducing working days, travel restrictions, quarantine and isolation), and development of interventions which can help to maintain the productivity of healthy population during the pandemic.

## Introduction

Severe acute respiratory syndrome coronavirus 2 (SARS-CoV-2), also known as coronavirus disease 2019 (COVID-19), is a novel betacoronavirus which was first identified in Wuhan in the Hubei province of China in December 2019. As with SARS-CoV, SARS-CoV-2 causes fever, cough, shortness of breath, pneumonia and lung infections, and results in high morbidity and mortality. Since January 2020, COVID-19 has rapidly spread through China and worldwide. Until the end of March 2020, 750,890 confirmed cases were identified worldwide, of which 82,545 cases were from China.^1^

Prevention and treatment of COVID-19 can be expensive. According to the Chinese clinical guidelines,^2,3^ all confirmed cases with COVID-19 should receive inpatient care. Moreover, patients with critical COVID-19 often require expensive medical procedures such as use of mechanical ventilation and extracorporeal membrane oxygenation (ECMO). These could potentially pose a substantial economic burden on the healthcare system. The societal cost of COVID-19 could be even larger. In order to prevent disease transmission, a series of emergency measures were implemented by the Chinese government,^4^ including isolation of suspected and confirmed cases, 14-day quarantine for close contacts of suspected or confirmed cases, lock down of Wuhan city and the adjacent areas, travel restrictions, school closure, shutting down of non-essential buildings, and extending the Chinese New Year holiday period. In order to delay the travel rush after the Chinese New Year holiday, the State Council extended the holiday period from 31^st^ January 2020 to 3^rd^ February 2020.^5^ On top on this, the local governments introduced their own policies to further extend the holiday. Whilst these containment strategies successfully reduced the transmission of COVID-19,^6^ they inevitably caused enormous productivity loss.

To our knowledge, no studies have quantified the cost-of-illness (COI) of COVID-19. This study aims to fill the gap by assessing the health and societal cost of the recent CVOID-19 outbreak in 31 provincial-level administrative regions in mainland China.

## Methods

This study was conducted and reported in accordance with the COI evaluation checklist developed by Larg *et al*.^7^

### Study population

The population of interest are all residents in mainland China, which consists of 31 provincial-level administrative regions, including 23 provinces, five autonomous regions (Inner Mongolia, Guangxi Zhuang, Tibet, Ningxia Hui and Xinjiang Uyghur) and four municipalities (Shanghai, Beijing, Tianjin and Chongqing). The population of interest was divided into four mutually exclusive patient subgroups, based on their experience of COVID-19: (i) asymptomatic close contacts of suspected or confirmed cases, who were eventually diagnosed as COVID-19 negative; (ii) symptomatic suspected cases with or without close contact history with existing suspected cases or confirmed cases, who were eventually diagnosed as COVID-19 negative; (iii) confirmed cases, including those previously assessed as close contacts or suspected cases; and (iv) people not considered to have had contact with COVID-19. Confirmed cases were further divided into non-severe, severe and critical cases, according to the disease severity. The definitions of each patient subgroup are reported in detail in the Appendix, Section 1. The diagnostic and treatment pathway for each patient subgroup is illustrated in Figure 1, and described in the Appendix, Section 2.

**Figure 1:**
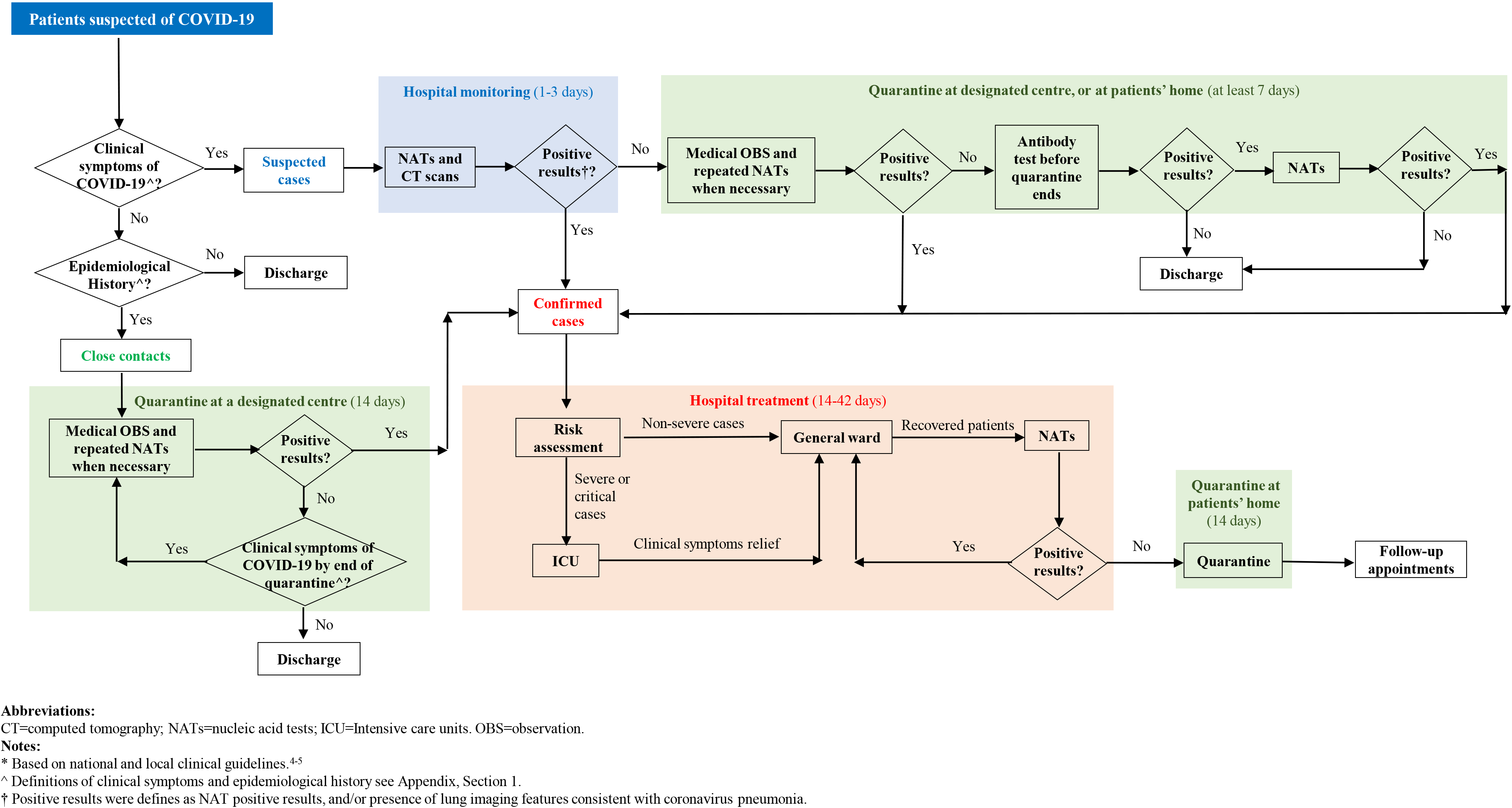
Simplified diagnostic and treatment pathway for COVID-19*

### Study period

Although COVID-19 was first identified in China in December 2019, the majority of confirmed cases (99.97%) were identified in 2020.^8^ Therefore, the costing period was set from 1^st^ January 2020 to 31^st^ March 2020.

### Perspective and outcomes

Two costing perspectives were adopted: healthcare and societal. From the healthcare perspective, the cost of both routine healthcare and non-routine healthcare were included. Routine healthcare includes identification, diagnosis, treatment and follow-up for people suspected of, or with a diagnosis of COVID-19. Non-routine healthcare includes (i) risk subsidy for front-line healthcare professionals who work with suspected and/or confirmed cases; and (ii) emergency funds for construction of temporary emergency buildings (i.e. the Huoshenshan hospital, the Leishenshan hospital, and the mobile cabin hospital in Wuhan city), and for emergency procurement of medical supplies and equipment (as opposed to routine procurement). From the societal perspective, the cost components included direct healthcare costs (as described above), direct nonhealthcare cost (compulsory quarantine for close contacts and suspected cases),^2^ and productivity losses. The quarantine cost can be covered by the local government, or by the quarantined individual, or jointly, depending on local policies. The outcomes of interest were total healthcare and societal cost for each region, and for mainland China. All costs were expressed in RMB (2019 value) and converted to USD using the OECD annual exchange rate for 2019 (1 USD = 6.91 RMB).^9^

### Methods for cost estimation

There are two broad approaches for estimating cost in COI: the bottom-up approach and the top-down approach.^7^ The bottom-up approach assesses the average cost per patient and multiplies it by the prevalence of the illness. The top-down approach uses aggregated data along with a population-attributable fraction to assign a percentage of total expenditures (the attributable costs) to the disease of interest. Both approaches are defensible; there is no clear consensus on which approach is more appropriate than the other. In this study, the feasibility of using both approaches were explored. Due to a lack of published total expenditures on COVID-19 (details see the Appendix, Section 3), a bottom-up approach was adopted for this study.

#### Epidemiological data

The National Health Commission (NHC) of the People’s Republic of China publishes daily updates of the COVID-19 statistics.^10^ However, it only reports the national statistics without detailed information for each region, except Hubei province. Therefore, the number of newly identified close contacts, suspected cases and confirmed cases in each region was manually extracted from the daily updates reported by the local health commission of each region (for data sources see Appendix, Table 1). While all regions reported complete data for the number of confirmed cases and the numbers of deaths of confirmed cases, not all of them reported complete data for the number of close contacts and/or suspected cases. In such cases, missing data were either estimated from the published literature, or calibrated from the reported national number of confirmed cases,^10^ assuming the same underlying ratio between the number of close contacts/suspected cases and confirmed cases across regions. It was estimated that during the study period, there were 707,913 close contacts, 126,032 suspected cases and 81,879 confirmed cases in mainland China. 5.2% of close contacts and 65.0% of suspected cases were diagnosed as COVID-19. Of those confirmed cases, 80.2% were non-severe cases, 13.6% were severe cases and 6.2% were critical cases. 83.2% of confirmed cases were from Hubei province. The detailed estimation methods and results by region are reported in the Appendix, Section 4.

#### Direct healthcare cost

The healthcare resource use for close contacts, suspected cases and confirmed cases were informed by published literature^11,12^ and clinical guidelines,^2,3^ supplemented with expert opinion where there was a lack of data. Shanghai is one of the few regions in China which reports comprehensive unit cost data.^13^ In order to calculate the unit costs for other regions, an ‘Healthcare industry salary index’ was calculated to estimate unit costs for each region (Appendix, Section 5), by dividing the average healthcare industry salary in the region of interest, by the average healthcare industry salary in Shanghai.^14^ Table 1 presents the healthcare cost per person for each patient subgroup, based on the estimated resource use and weighted national unit cost calculated based on the ‘Healthcare industry salary index’. The healthcare cost of managing close contacts diagnosed as COVID-19 negative, and suspected cases diagnosed as COVID-19 negative was 584 RMB (85 USD) and 974 RMB (141 USD) per person, respectively. The average cost of treating a confirmed case was 22,355 RMB (3,235 USD), ranging from 6,489 RMB (939 USD) for non-severe cases to 176,744 RMB (25,578 USD) for critical cases. The detailed methods for estimating healthcare cost for each patient subgroup and results by region are reported in the Appendix, Section 6.1–6.2.

**Table 1:**
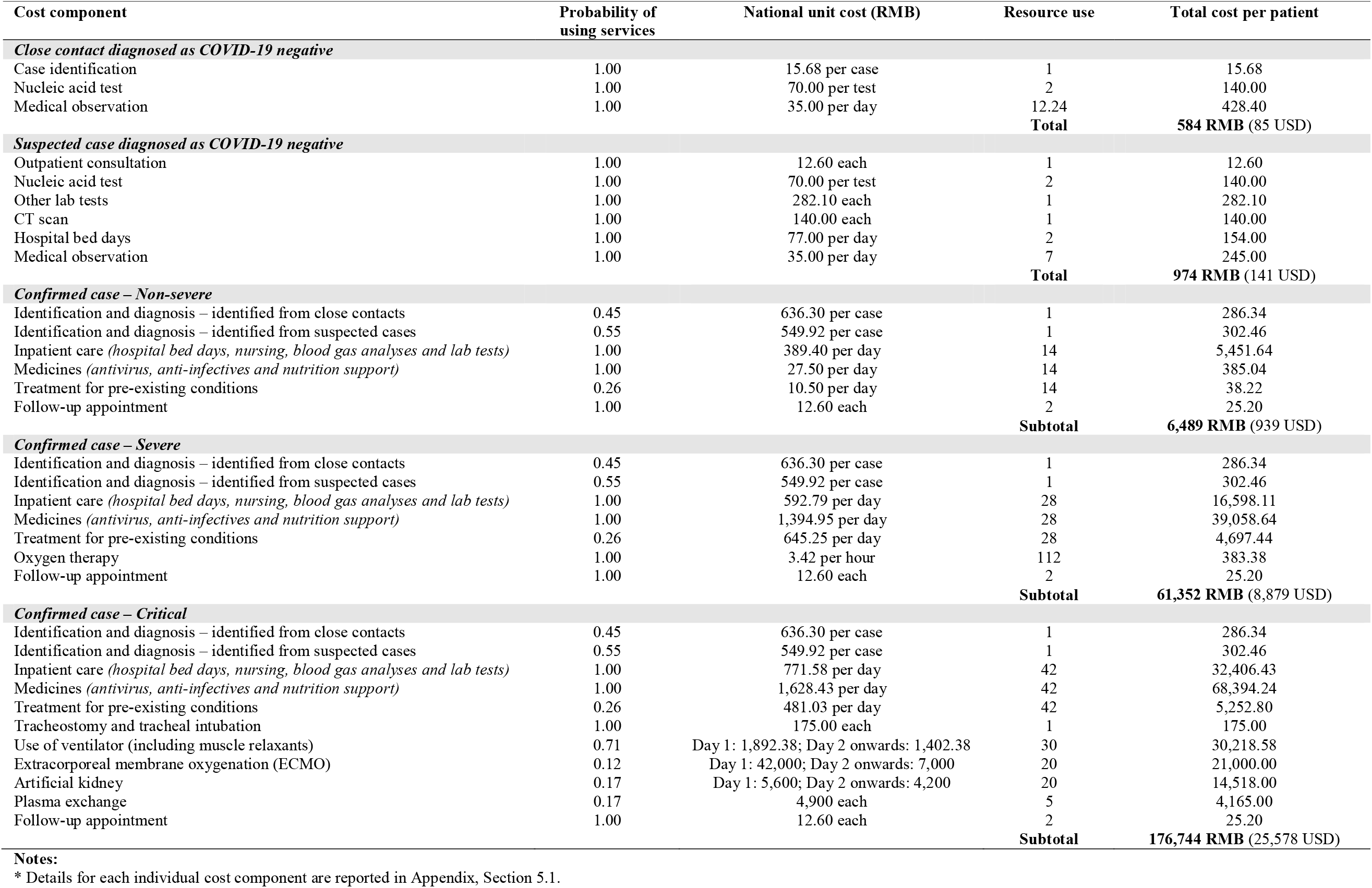
Summary of healthcare cost for close contact, suspected case and confirmed case (per person) *^*^*.

| Cost component | Probability of using services | National unit cost (RMB) | Resource use | Total cost per patient |
| --- | --- | --- | --- | --- |
| **Close contact diagnosed as COVID-19 negative** |  |  |  |  |
| Case identification | 1.00 | 15.68 per case | 1 | 15.68 |
| Nucleic acid test | 1.00 | 70.00 per test | 2 | 140.00 |
| Medical observation | 1.00 | 35.00 per day | 12.24 | 428.40 |
|  |  |  | **Total** | **584 RMB (85 USD)** |
| **Suspected case diagnosed as COVID-19 negative** |  |  |  |  |
| Outpatient consultation | 1.00 | 12.60 each | 1 | 12.60 |
| Nucleic acid test | 1.00 | 70.00 per test | 2 | 140.00 |
| Other lab tests | 1.00 | 282.10 each | 1 | 282.10 |
| CT scan | 1.00 | 140.00 each | 1 | 140.00 |
| Hospital bed days | 1.00 | 77.00 per day | 2 | 154.00 |
| Medical observation | 1.00 | 35.00 per day | 7 | 245.00 |
|  |  |  | **Total** | **974 RMB (141 USD)** |
| **Confirmed case – Non-severe** |  |  |  |  |
| Identification and diagnosis – identified from close contacts | 0.45 | 636.30 per case | 1 | 286.34 |
| Identification and diagnosis – identified from suspected cases | 0.55 | 549.92 per case | 1 | 302.46 |
| Inpatient care (*hospital bed days, nursing, blood gas analyses and lab tests*) | 1.00 | 389.40 per day | 14 | 5,451.64 |
| Medicines (*antivirus, anti-infectives and nutrition support*) | 1.00 | 27.50 per day | 14 | 385.04 |
| Treatment for pre-existing conditions | 0.26 | 10.50 per day | 14 | 38.22 |
| Follow-up appointment | 1.00 | 12.60 each | 2 | 25.20 |
|  |  |  | **Subtotal** | **6,489 RMB (939 USD)** |
| **Confirmed case – Severe** |  |  |  |  |
| Identification and diagnosis – identified from close contacts | 0.45 | 636.30 per case | 1 | 286.34 |
| Identification and diagnosis – identified from suspected cases | 0.55 | 549.92 per case | 1 | 302.46 |
| Inpatient care (*hospital bed days, nursing, blood gas analyses and lab tests*) | 1.00 | 592.79 per day | 28 | 16,598.11 |
| Medicines (*antivirus, anti-infectives and nutrition support*) | 1.00 | 1,394.95 per day | 28 | 39,058.64 |
| Treatment for pre-existing conditions | 0.26 | 645.25 per day | 28 | 4,697.44 |
| Oxygen therapy | 1.00 | 3.42 per hour | 112 | 383.38 |
| Follow-up appointment | 1.00 | 12.60 each | 2 | 25.20 |
|  |  |  | **Subtotal** | **61,352 RMB (8,879 USD)** |
| **Confirmed case – Critical** |  |  |  |  |
| Identification and diagnosis – identified from close contacts | 0.45 | 636.30 per case | 1 | 286.34 |
| Identification and diagnosis – identified from suspected cases | 0.55 | 549.92 per case | 1 | 302.46 |
| Inpatient care (*hospital bed days, nursing, blood gas analyses and lab tests*) | 1.00 | 771.58 per day | 42 | 32,406.43 |
| Medicines (*antivirus, anti-infectives and nutrition support*) | 1.00 | 1,628.43 per day | 42 | 68,394.24 |
| Treatment for pre-existing conditions | 0.26 | 481.03 per day | 42 | 5,252.80 |
| Tracheostomy and tracheal intubation | 1.00 | 175.00 each | 1 | 175.00 |
| Use of ventilator (including muscle relaxants) | 0.71 | Day 1: 1,892.38; Day 2 onwards: 1,402.38 | 30 | 30,218.58 |
| Extracorporeal membrane oxygenation (ECMO) | 0.12 | Day 1: 42,000; Day 2 onwards: 7,000 | 20 | 21,000.00 |
| Artificial kidney | 0.17 | Day 1: 5,600; Day 2 onwards: 4,200 | 20 | 14,518.00 |
| Plasma exchange | 0.17 | 4,900 each | 5 | 4,165.00 |
| Follow-up appointment | 1.00 | 12.60 each | 2 | 25.20 |
|  |  |  | **Subtotal** | **176,744 RMB (25,578 USD)** |

**Notes:**
\* Details for each individual cost component are reported in Appendix, Section 5.1.

According to the State Council,^15^ there were 42,600 front-line health professionals working with suspected and/or confirmed cases. The daily risk subsidy for front-line healthcare professionals was estimated to be 300 RMB per person. The emergency funds were estimated based on the budget plans of the Ministry of Finance and the National Development and Reform Commission. For reusable equipment, only the cost attributable to the three-month period was included in our analysis. The calculation methods and results for emergency funds are reported in the Appendix, Section 6.3.

#### Direct non-healthcare cost

The average cost of quarantine for close contacts and suspected cases was estimated at 1,246 RMB (180 USD) and 735 RMB (106 USD) per person, based on national clinical guidelines^2^ and expert opinion. The detailed methods and results for calculating quarantine cost are reported in Appendix, Section 7.

#### Productivity loss

Under the bottom-up approach, there are two primary methods to estimate productivity loss: the human capital approach (HCA) and the friction cost approach (FCA).^16^ The HCA captures lost productivity over the entire period of lost working opportunity, by multiplying the earnings lost for different age and gender groups by the corresponding number of people in that group. The FCA is similar to HCA, but assumes that people who stop working due to illness will be eventually be replaced by someone who was previously unemployed, and therefore only measures the productivity losses during the time it takes to replace a worker.^17^ There is no consensus on which approach is superior to the others For this study, FCA was considered to be inappropriate as it was difficult to find any replacement workers during the COVID-19 outbreak. Therefore, HCA was used to estimate the productivity loss for this study.

According to the China Statistical Yearbook,^14^ there were 416.5 million working people in mainland China in Year 2018. The top three regions with the highest number of working people are Guangdong province (13.9%), Jiangsu province (10.1%) and Zhejiang province (6.7%). The national average daily salary is 271 RMB (39 USD), ranging from 204.7 RMB (29.6 USD) in Heilongjiang province to 486.4 RMB (70.4 USD) in Beijing. The national unemployment rate is 3.0%, ranging from 1.4% in Beijing to 4.0% in Heilongjiang province. There was a lack of data on the employment status for close contacts, suspected cases and confirmed cases. Therefore, the employment rate for each patient subgroup was estimated at 55% based on the age and sex distribution of confirmed cases, the legal working age (16 years old) and the official retirement age (60 years old for men and 50 years old for women), and the national unemployment rate (3.0%). The average working days lost for people not considered to having contracted COVID-19 was estimated at 23.26 days, based on “Baidu Migration Index”,^18^ which tracks the proportion of workers returning from their hometowns to work after the Chinese New Year holiday. This index is calculated by Baidu, Inc, using artificial intelligence based on data collected from over 348 million active users of Baidu Maps.^19^ Close contacts, suspected cases and confirmed cases may have experienced more working days lost due to their quarantine and/or hospitalisation.^11,12,20^ Working days lost for people in this group depended on the start and end date of their quarantine and/or hospitalisation, and whether these overlapped with (i) the extended Chinese New Year holiday; and (ii) the study period. The detailed calculation process for estimating number of working people and length of the working days lost for each patient subgroup are reported in Appendix, Section 8.

### Sensitivity analysis

We identified costs which contributed to 10% or more of the total health care costs and societal costs and we varied the parameters for resource use and unit cost which underpinned these costs in sensitivity analysis. The ranges for the selected parameters were informed by available data or by judgement.

## Results

Costs are reported in Table 2. The routine healthcare cost was estimated at 2.16 billion RMB (0.32 billion USD). The top three cost components of routine healthcare cost were: inpatient care (41.0%), medicines (30.9%) and medical observation for close contacts and suspected cases (12.4%). In terms of population, critical confirmed cases consumed most routine healthcare cost (34.3%), followed by severe confirmed cases (26.4%), close contacts diagnosed as COVID-19 negative (16.7%) and non-severe confirmed cases (15.7%). The non-routine healthcare cost was estimated to be 2.10 billion RMB (0.30 billion USD), of which 40.0% was risk subsidy for front-line healthcare professionals, and 60.0% was emergency funds. The total societal cost of COVID-19 was estimated at 2,647 billion RMB (383 billion USD), which is equivalent to 2.7% of China’s gross domestic product (GDP) in 2019 ($14.14 trillion USD).^21^ Healthcare costs accounted for only 0.2% of the societal cost while productivity losses accounted for 99.8%. Productivity losses were almost entirely attributable to people not diagnosed with COVID-19 who experienced lost working time due to the government policies controlling population movement (99.7%). The healthcare cost and societal cost for each region are shown in Figure 2 and 3, respectively. The healthcare cost for Hubei province accounted for 83.2% of the national healthcare cost, which far exceeded the other regions. Guangdong province incurred the highest societal cost (14.6%), followed by Jiangsu province (10.1%) and Beijing (7.7%).

**Table 2:**
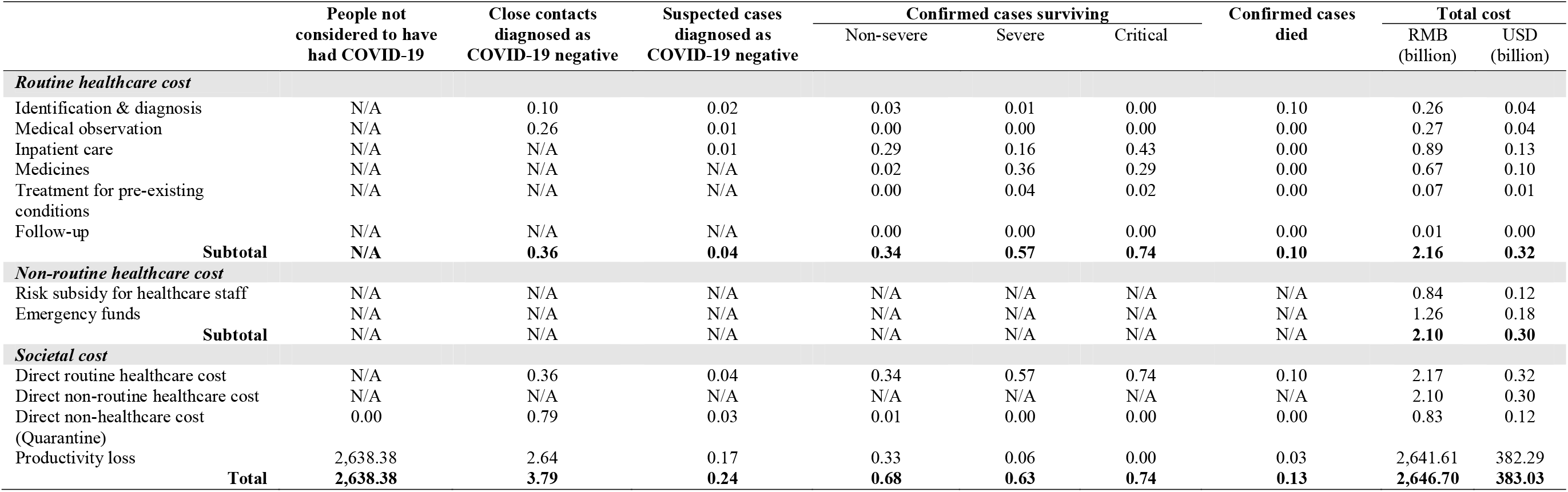
Breakdown of healthcare cost and societal cost estimated using bottom-up approach (RMB billion)

**Fig 3:**
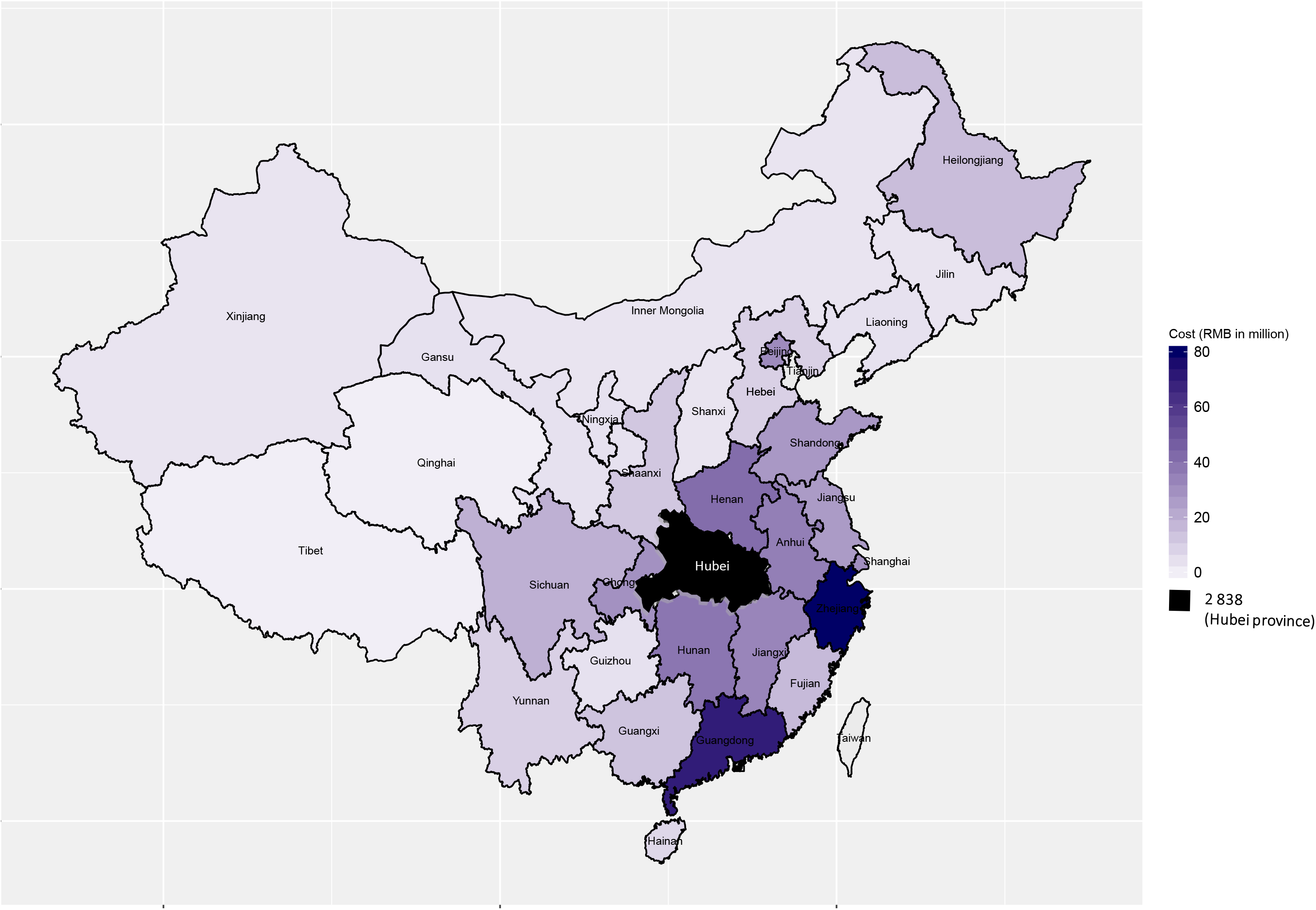
Healthcare cost of COVID-19 by region

The results of the sensitivity analyses for healthcare cost and societal cost are presented in Figure 4 and 5, respectively. The direct healthcare cost was most sensitive to the proportion of confirmed cases with severe or critical disease, and the healthcare cost per person for managing severe and critical confirmed cases. The productivity loss was most sensitive to the number of working days lost for people not considered to have had COVID-19, and the national average daily salary.

**Figure 4:**
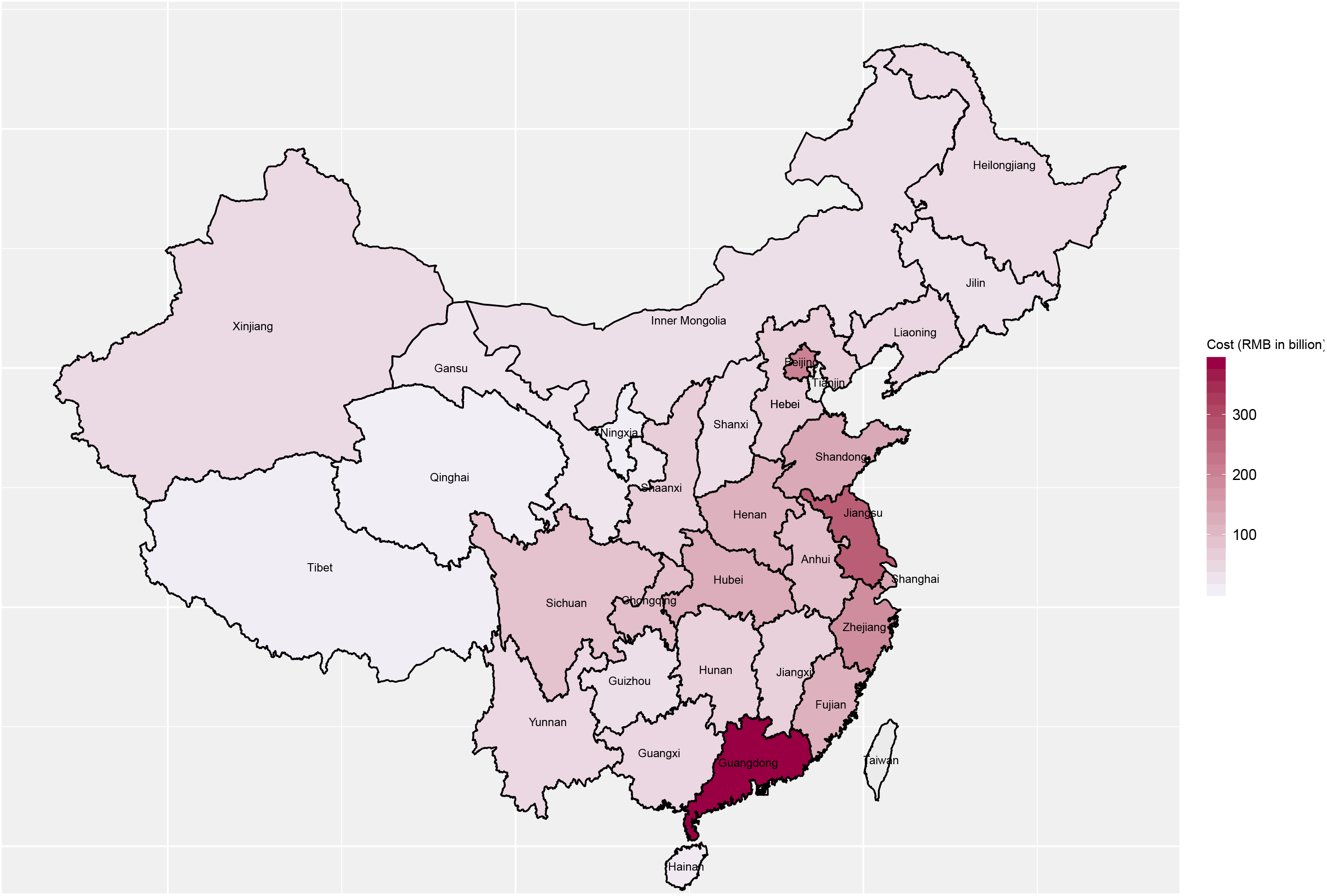
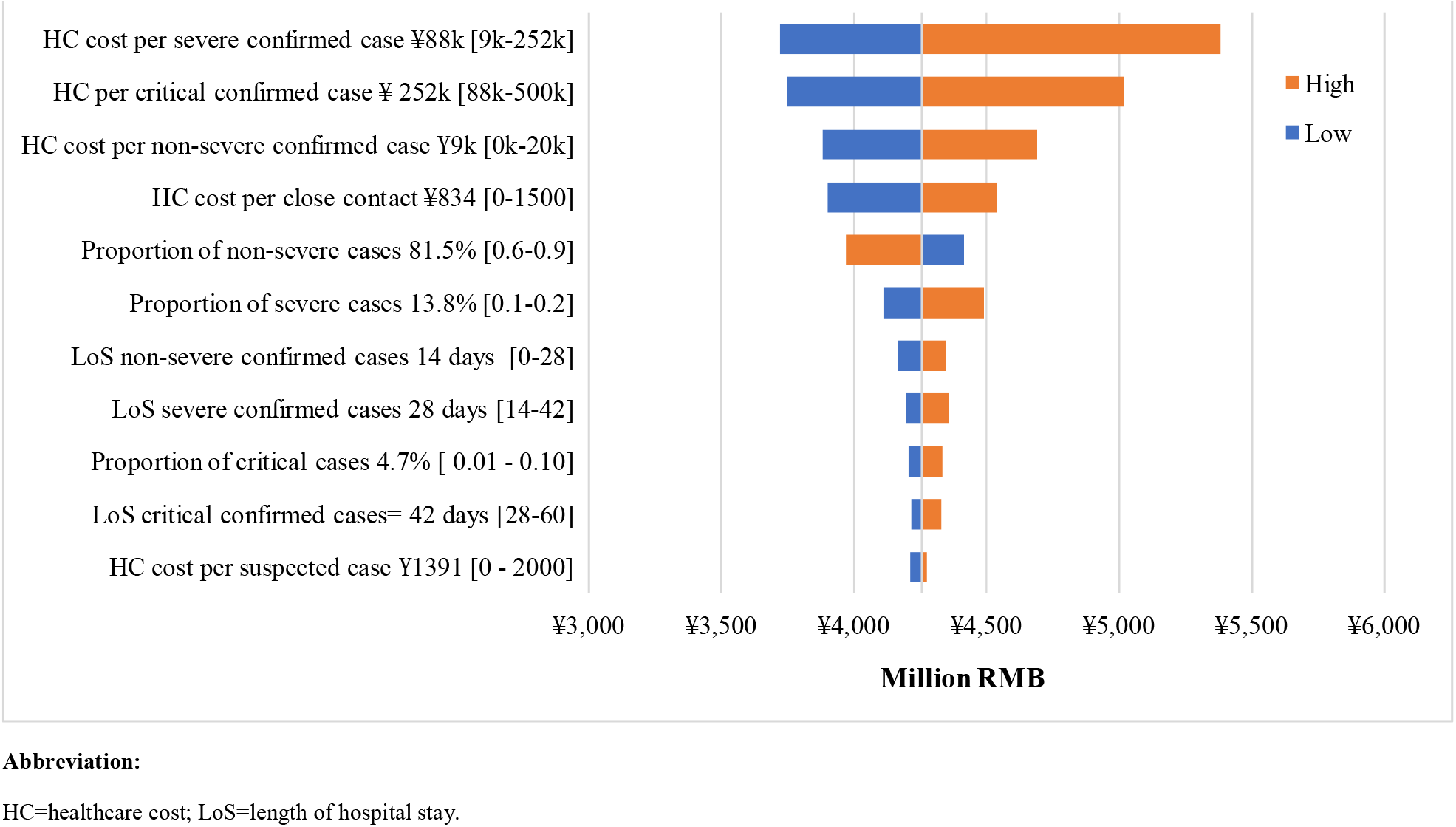
Sensitivity analysis results for healthcare cost

**Figure 5:**
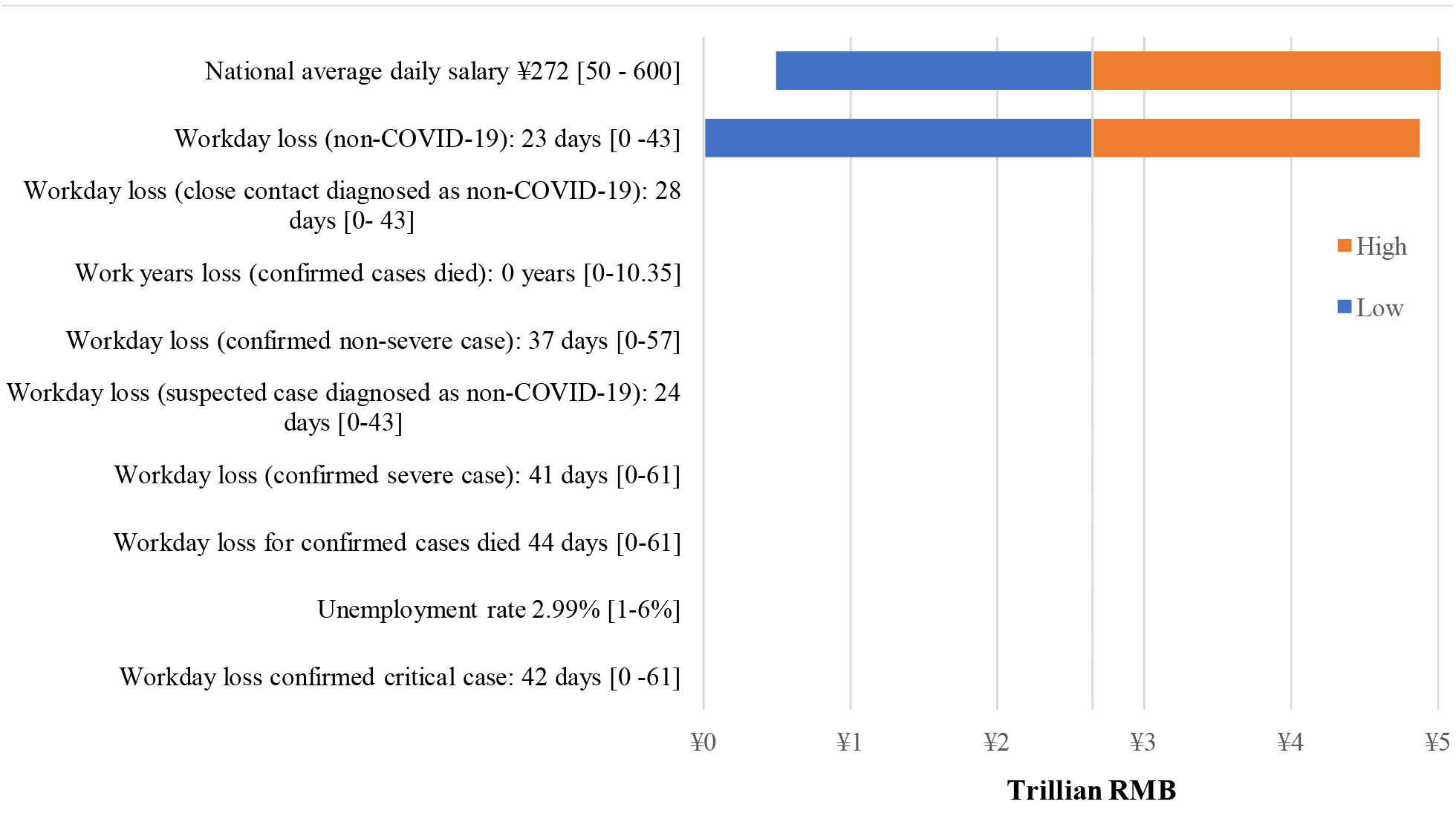
Sensitivity analysis results for societal cost

## Discussion

Our study estimated the healthcare and societal cost associated with the COVID-19 outbreak for the first three months of 2020 at 0.62 billion USD and 383 billion USD, respectively. Although the routine healthcare cost per person for confirmed cases was high (ranging from 939 USD for non-severe cases to 25,578 USD for severe cases), 99.7% of the societal cost was attributable to productivity losses in people not considered to have had COVID-19. This reflects the overall number of employed people (416.5 million), which is much larger than the number of confirmed cases (80,000).

Hubei province, where the majority of confirmed cases were identified, accounted for 83.2% of the national healthcare cost. The productivity loss was greatest for those regions with the highest number of employed people and/or with highest daily salary, such as Guangdong province (57.7 million employed people, 296 RMB), Jiangsu province (42.2 million employed people, 279 RMB) and Beijing (15.7 million employed people, 486 RMB).

### Comparison of results with published data/literature

Costs from the literature are inflated to 2019 values using a local Consumer Price Index, and converted to USD using the OECD annual exchange rate to aid comparison.^22^ Our rapid review did not identify any existing COI studies for COVID-19. However, several studies have examined the cost of infectious respiratory diseases caused by other types of coronavirus.^22^ The additional healthcare cost of implementing infection prevention measures for Middle East respiratory syndrome (MERS) in a UK hospital was reported at $165 USD per day.^23^ This study did not include the treatment cost of MERS as all patients with suspected MERS were eventually diagnosed with influenza. Three studies^24–26^ reported the cost of managing patients with SARS and preventing transmission in a single hospital based on retrospective analysis of financial records. The reported healthcare cost per SARS case ranged from 4,151 USD in mainland China^25^ to 362,700 USD in Canada.^24^ The cost for mainland China is comparable to our estimate of 3,235 USD per COVID-19 case.^25^ Two studies conducted in Taiwan compared the resource use for all medical conditions before and after outbreak of SARS based on retrospective analysis of database from a single hospital^27^ and the national health insurance data.^28^ Both studies reported a reduction in healthcare resource use during the peak of the SARS epidemic, including emergency department visits,^27,28^ inpatient care and dental care.^28^ The authors assumed that the reduction in resource use was probably caused by people’s fears of SARS.^18,19^ Resource use gradually returned to normal levels a few months after the SARS epidemic was over. Neither studies assessed the cost impact of SARS beyond a year. Pan and Liu^29^ analysed Chinese governmental health expenditure (GHE) during 2002–2006, and found that the SARS outbreak in 2003 increased GHE by 4.1%, which is equivalent to 0.20% of China’s GDP in 2013.

Our estimated productivity loss (382 billion USD) was comparable to the decrease in Chinese GDP for the first quarter of 2020 compared to the same period in 2019 (218 billion USD).^21^ Lee and McKibbin^30^ used a simulation model to estimate the societal cost of SARS in 30 countries. They estimated the societal cost of SARS in mainland China to be 1.05% of China’s GDP,^30^ which is to the same order of magnitude as our estimate of the societal cost of COVID-19 (2.7% of China’s GDP in 2019^21^).

### Implications for policy and future research

The societal cost of COVID-19 is enormous and vastly outweighs the health care cost. In the absence of evidence on cost-effectiveness countries have shown a willingness to impose unprecedented controls on citizens’ movements and ability to work in order to combat the spread of Covid-19. Future work will examine the cost-effectiveness of these policies. Our data can help inform these analyses. Currently, the path to the end of the Covid-19 epidemic is unclear. However, as the world emerges from this pandemic a conversation will begin on the appropriate measures to mitigate the impact of future emerging diseases. Our analysis underlines the importance of collective action to strengthen health systems, particularly the capacity to test for infection and trace contacts, and the capacity to treat patients with severe lung infections. Effective action will require international cooperation and significant investment. Our analysis, which demonstrates the extent of the impact of Covid-19 beyond the health care system, justifies the redirection of resources from other sectors of the economy to strengthen health systems. Without significant additional resources, underinvestment in the capacity of health systems to combat future pandemics could prove to be far costlier than the additional investment required.

### Strengths and limitations

#### Strengths

There are a number of strengths of this study. Firstly, this study fills an important evidence gap by presenting the first COI study of COVID-19. The study identifies the magnitude of the cost impact on different sectors of the economy which is necessary to inform planning of services and the prioritisation of research. The data reported in this study also provide important information for future economic evaluations of interventions (e.g. cost-effectiveness studies) for COVID-19. Our study accessed detailed resource use data across the 31 regions of mainland China, including obtaining the incidence data from the local health commission of each region. We applied unit cost data adjusted to reflect relative price differentials across provinces, and benefitted from expert clinician (SY from Shanghai and SZ from Hubei province) to check the resource use for each patient subgroup. Productivity costs for close contacts, suspected cases and confirmed cases were carefully estimated based on the duration of quarantine and/or treatment, and regional migration patterns following the end of the extended New Year holiday period.

#### Limitations

Our analysis is subject to some limitations. Firstly, our analysis extends only to the first three months of the epidemic. There are likely to be longer term economic impacts (e.g. reduced international trade and increased unemployment rates) that were not captured. Second, due to a lack of data, some cost components were not included, such as productivity losses for carers of suspected/confirmed cases and out-of-pocket payments for travel to hospitals and over-the-counter medicines. Third, due to a shortage of nucleic acid tests in China in January 2020, not all suspected patients received nucleic acid tests.^3^ Therefore, the reported number of confirmed cases is likely to be an underestimate, especially in Hubei province. Fourth, our estimate of the number of working days lost, which was based on migration data. may have overestimated losses for those people who worked from home. Finally, we lacked some data on the incidence, demographic information and prognosis for close contacts and suspected cases, and had to estimate them based on published literature and/or expert opinion.

## Conclusion

This paper presents the first COI study which quantifies the health and societal cost of COVID-19 in China. Evidence indicates that the control measures to prevent the spread of disease in the first three months of this year incurred substantial productivity costs amounting to over 2% of China’s annual GDP. These costs dwarfed the direct health care costs. The results of this study highlight the substantial economic burden of COVID-19 outbreak and provide evidence to justify significant investment in prevention and control measures for future outbreaks.

## Data Availability

All data used to support the analysis have been reported in either the main paper or the appendix.

## Acknowledgments

None.

## Compliance with Ethical Standards

### Contributors

HJ, HW and MP conceived the research idea. HJ and XL conducted literature review of existing COI studies of COVID-19. HW, PZ and JZ collected input data in China. SY and SZ advised on the resource use of preventing and treating COVID-19. HJ and HW conducted cost calculation. XL and MP checked input data and cost calculation. HJ, HW and XL wrote the first draft of the paper, which was subsequently been edited by all authors who have approved the final version. HJ and HW contributed equally to this work. HJ will serve as a guarantor for the overall content of the manuscript.

## Contract details of corresponding author

Name: Dr Huajie Jin, Tel: +44 (0)20 7848 0878, Address: King’s Health Economics, Institute of Psychiatry, Psychology & Neuroscience at King’s College London Box, 024, The David Goldberg Centre, De Crespigny Park, Denmark Hill, London SE5 8AF, UK

## Reference

1. World Health Organisation. Coronavirus disease 2019 (COVID-19) Situation Report – 712020. https://www.who.int/docs/default-source/coronaviruse/situation-reports/20200331-sitrep-71-covid-19.pdf?sfvrsn=4360e92b_4 (accessed 07/04/2020).

2. The National Health Commission of the People’s Republic of China. Guidance on infection prevention and control for COVID-19 (7th edition), 2020. http://www.gov.cn/zhengce/zhengceku/2020-03/04/5486705/files/ae61004f930d47598711a0d4cbf874a9.pdf (accessed 09/04/2020).

3. First Affiliated Hospital of Zhejiang University School of Medicine. Handbook of COVID-19 Prevention and Treatment, 2020. https://covid-19.alibabacloud.com/ (accessed 10/04/2020).

4. Muhammad F. China combating COVID-2019: lessons for unprepared South Asia, 2020. https://www.fudan.edu.cn/en/2020/0306/c1092a104273/page.htm (accessed 07/04/2020).

5. General Office of the State Council. General Office of the State Council’s announcement of extending the Chinese New Year holiday, 2020. http://www.gov.cn/zhengce/content/2020-01/27/content_5472352.htm (accessed 07/04/2020).

6. Zhang J, Litvinova M, Wang W, et al. Evolving epidemiology and transmission dynamics of coronavirus disease 2019 outside Hubei province, China: a descriptive and modelling study. The Lancet Infectious diseases 2020.

7. Larg A, Moss JR. Cost-of-illness studies: a guide to critical evaluation. Pharmacoeconomics 2011; 29(8): 653–71.

8. Wu Z, McGoogan, JM. Characteristics of and Important Lessons From the Coronavirus Disease 2019 (COVID-19) Outbreak in China: Summary of a Report of 72H314 Cases From the Chinese Center for Disease Control and Prevention. JAMA 2020.

9. OECD. Exchange rates, 2020. https://data.oecd.org/conversion/exchange-rates.htm (accessed 08/04/2020).

10. The National Health Commission of the People’s Republic of China. The latest update of COVID-19 in China, 2020. http://www.nhc.gov.cn/xcs/yqtb/list_gzbd.shtml (accessed 02/04/2020).

11. World Health Organisation. Report of the WHO-China Joint Mission on Coronavirus Disease 2019 (COVID-19), 2020. https://www.who.int/docs/default-source/coronaviruse/who-china-jointmission-on-covid-19-final-report.pdf (accessed 02/04/2020).

12. Cheng K, Wei M, Shen H. The clinical characteristics of minor and severe cases of 463 patients recovered from COVID-19,. Shanghai Medicine 2020: 1–15.

13. The Health Commission of Shanghai. Price of healthcare services provided by health care providers in Shanghai, 2020. http://wsjkw.sh.gov.cn/ylsfbz/index.html (accessed 03/04/2020).

14. National Bureau of Statistics of China. China Statistical Yearbook 2019. Beijing, China: China Statistics Press; 2020.

15. The State Council. Press Conference of the Joint Prevention and Control Mechanism of the State Council – 8th March 2020. Beijing, P.R. China; 2020.

16. Segel JE. Cost-of-illness studies—A primer. RTI-UNC Center of Excellence in Health Promotion Economics RTI international 2006; 1–39.

17. Koopmanschap MA, Rutten FF, van Ineveld BM, van Roijen L. The friction cost method for measuring indirect costs of disease. Journal of Health Economics 1995; 14(2): 171–89.

18. Baidu. Baidu Migration Index – 24th March 2020, 2020. https://mp.weixin.qq.com/s/zn4qME7XGSwMcfhufnpXeA (accessed 16/04/2020).

19. Smith C. 90 Amazing Baidu Statistics and Facts (2020) | By the Numbers, 2019. https://expandedramblings.com/index.php/baidu-stats/ (accessed 09/04/2020).

20. The People’s Government of Shanghai. Human Resources and Social Affairs Bureau’s response to delay in work resumption, 2020. http://www.shanghai.gov.cn/nw2/nw2314/nw32419/nw48516/nw48545/u26aw63619.html (accessed 05/04/2020).

21. National Bureau of Statistics of China. Gross domestic product in China, 2020. http://www.stats.gov.cn/tjsj/ (accessed 17/04/2020).

22. OECD. Consumer price indices (CPIs) – Complete database, 2020. https://stats.oecd.org/Index.aspx?DataSetCode=PRICES_CPI (accessed 08/04/2020).

23. Veater J, Wong N, Stephenson I, et al. Resource impact of managing suspected Middle East respiratory syndrome patients in a UK teaching hospital. Journal of Hospital Infection 2017; 95(3): 280–5.

24. Achonu C, Laporte A, Gardam MA. The financial impact of controlling a respiratory virus outbreak in a teaching hospital: lessons learned from SARS. Canadian Journal of Public Health 2005; 96(1): 52–4.

25. Xiao F, Chen BW, Wu YF, et al. [Analysis on the cost and its related factors of clinically confirmeds severe acute respiratory syndrome cases in Beijing]. Chung-Hua Liu Hsing Ping Hsueh Tsa Chih Chinese Journal of Epidemiology 2004; 25(4): 312–6.

26. Yazdanpanah Y, Daval A, Alfandari S, et al. Analysis of costs attributable to an outbreak of severe acute respiratory syndrome at a French hospital. Infection Control and Hospital Epidemiology 2006; 27(11): 1282–5.

27. Huang H, Yen H, Kao W, Wang L, Huang Chun I, Lee C. Declining emergency department visits and costs during the severe acute respiratory syndrome (SARS) outbreak. Journal of the Formosan Medical Association 2006; 105(1): 31–7.

28. Chang HJ, Huang N, Lee CH, Hsu YJ, Hsieh CJ, Chou YJ. The impact of the SARS epidemic on the utilization of medical services: SARS and the fear of SARS. American Journal of Public Health 2004; 94(4): 562–4.

29. Pan J, Liu GG. The determinants of Chinese provincial government health expenditures: evidence from 2002–2006 data. Health Economics 2012; 21(7): 757–77.

30. Lee J, McKibbin, W. Learning from SARS. Preparing for the Next Disease Outbreak. In: Knobler S, Mahmoud A, Lemon S, Mack A, Sivitz L, Oberholtzer K, editors. Washington (DC): National Academies Press (US); 2004.

